# Development and pilot testing of an empowerment-based intervention for adolescents and young adults living with HIV transitioning to adult HIV care in Uganda

**DOI:** 10.64898/2026.06.19.26355595

**Authors:** Scholastic Ashaba, Alain Favina, Charles Baguma, Patricia Tushemereirwe, Denis Nansera, Alison Comfort, Jessica M. Perkins, Samuel Maling, Brian C. Zanoni, Alexander C. Tsai

## Abstract

**Background:** Transition from adolescent to adult HIV care is a vulnerable period for adolescents and young people living with HIV (AYLHIV). It is marked by increasing responsibility for health management alongside ongoing psychosocial, developmental, and structural challenges. Many AYLHIV report limited preparation for adult care. Existing transition interventions often prioritize biomedical outcomes and are developed in high-income settings, with less emphasis on empowerment and contextual relevance in low– and middle-income countries. This study aimed to develop and assess the feasibility and acceptability of an empowerment-focused intervention to support AYLHIV during transition to adult HIV care.

**Methods:** We developed the Empowerment and Personal Transformation (EPT) intervention, a six-module psychosocial program designed to address communication, self-management, emotional regulation, and personal growth during transition. Intervention content was informed by qualitative data collected through in-depth interviews with AYLHIV and key stakeholders during the intervention development phase. The intervention was subsequently implemented, and feasibility and acceptability were assessed using quantitative measures. Internal consistency of feasibility and acceptability scales was evaluated using Cronbach’s alpha coefficients.

**Results:** The EPT intervention demonstrated high feasibility and acceptability among participants. The intervention was perceived as practical to deliver, not burdensome, and responsive to participants’ transition-related experiences. Acceptability findings indicated strong participant engagement and approval, with participants describing the intervention as relevant to their psychosocial needs, including experiences of stigma and challenges related to transitioning to independent HIV care. The acceptability subscale demonstrated excellent internal reliability (Cronbach’s alpha = 0.92), while the feasibility subscale showed good internal consistency (Cronbach’s alpha = 0.86).

**Conclusions:** Findings suggest that empowerment-focused interventions are feasible and acceptable for supporting AYLHIV transitioning to adult HIV care.

## Background

HIV remains a leading cause of death among adolescents despite widespread availability of antiretroviral medications (Mburu et al., 2014; Slogrove et al., 2017). Generally, adolescents and young adults living with HIV (AYLHIV) have poor outcomes with higher rates of loss to follow-up and mortality compared with other age groups (Auld et al., 2014; MacPherson et al., 2015).

Mortality rates and loss to follow-up are highest during the transition from adolescent HIV clinics to adult HIV care (Auld et al., 2014; Davies et al., 2017). Timely transition to adult HIV care is important for AYLHIV to access developmentally appropriate services and achieve optimal health outcomes (Mburu et al., 2014; Schwartz et al., 2011). In addition, the transition represents a sensitive period that requires adequate structural and psychosocial support to ensure continuity of care and long-term wellbeing (Cervia, 2013). Successful transition is more likely among AYLHIV who are adequately prepared to manage their clinical care as well as the psychosocial demands of adult clinic settings (Crowley et al., 2019). Preparation for transition to adult HIV care often includes skills-based training to enhance self-efficacy and structured educational support to address gaps in HIV-related knowledge, both of which have been associated with improved clinical and psychosocial outcomes following transition (Ashaba et al., 2024; Tanner et al., 2016).

Despite national and international guidelines increasingly advocating for structured and readiness-based transition processes, their implementation remains inconsistent across different settings (Fair et al., 2010). In many low– and middle-income countries, including Uganda, transition to adult HIV care is often determined by chronological age rather than by any assessment of individual readiness (Ashaba et al., 2023; Dahourou et al., 2017; Shimbre et al., 2024). Qualitative studies from Uganda indicate that many AYLHIV transition to adult HIV clinics without adequate preparation, often lacking a clear understanding of how adult services operate and feeling uncertain about managing appointments, medication refills, and communication with healthcare providers (Ashaba et al., 2023; Ashaba et al., 2022).

Effective transition should extend beyond logistical aspects of care transfer to addressing psychosocial aspects of care transfer, including emotional readiness, empowerment, and self-efficacy (Hussen et al., 2017). Additionally, the timing of transition to adult HIV care often coincides with other major life transitions for AYLHIV, including completion of schooling, entry into the workforce, evolving family responsibilities, and the formation of intimate relationships, which can further complicate sustained engagement in care (Ashaba et al., 2023; Jao et al., 2016). These concurrent life transitions occur alongside the intersecting physical, psychological, and social demands of adolescence and young adulthood, intensified further by pressures related to peer acceptance and HIV-related stigma, thereby creating additional barriers to successful transition to adult HIV care and independent health management (Lanyon et al., 2020; Machado et al., 2016). These factors underscore that transition to adult HIV care extends beyond a simple administrative transfer of services and represents a complex developmental process requiring AYLHIV to build the skills, confidence, and resilience needed to manage their treatment, communicate effectively with healthcare providers, and navigate ongoing emotional and social challenges (Jao et al., 2016; Mofenson & Cotton, 2013).

In response to these challenges, there has been growing interest in empowerment-based interventions to support young people with chronic health conditions, including HIV, who are attempting to transition successfully to adult care (Acuña Mora et al., 2019). Empowerment-based interventions, particularly those that emphasize active participation, skills development, and supportive relationships, have been associated with improved engagement in care, enhanced responsiveness to health care, and treatment outcomes following transition (Akama et al., 2023; Enane et al., 2018; Ferry et al., 2022; Mulawa et al., 2023). Through empowerment processes, individuals acquire knowledge and skills that enhance self-efficacy, support informed decision-making, and promote independence from caregivers and healthcare providers (McDonagh & Kelly, 2010; Sell et al., 2016). For AYLHIV, empowerment may specifically involve developing confidence to engage with adult care providers, strengthening self-management and problem-solving skills, and fostering a positive sense of identity beyond HIV status (Areri et al., 2022; Ashaba et al., 2024; Mutumba et al., 2019; Ness et al., 2021).

Despite growing interest in empowerment-based approaches, evidence remains limited on how such interventions can be effectively designed and implemented to support transition to adult HIV care, particularly in resource-constrained settings. Many existing interventions have been developed in high-income settings or focus primarily on short-term outcomes, often failing to adequately address the practical constraints and health system realities of low– and middle-income countries (Aantjes et al., 2014; Areri et al., 2022). Furthermore, few studies have examined the acceptability of empowerment-based transition interventions from the perspectives of young people or their feasibility within routine HIV care settings (Garofalo et al., 2018; Kuhns et al., 2017). To address this gap in literature, we developed an empowerment and personal transformation (EPT) intervention for AYLHIV transitioning to adult HIV care in Uganda. In this manuscript we describe the development of the EPT intervention and report findings on its feasibility and acceptability among AYLHIV.

## METHODS

### Study design and theoretical framework

We employed a qualitative, participatory intervention development approach followed by pilot testing of the EPT intervention to assess its feasibility and acceptability among AYLHIV. The study was conducted in three phases: (1) a qualitative needs assessment to identify psychosocial and behavioral needs related to transition to adult HIV care; (2) intervention development informed by qualitative findings, theory, and stakeholder input; and (3) pilot implementation and evaluation of the intervention’s feasibility and acceptability. The development of the EPT intervention was informed by empowerment theory (Zimmerman, 2000) and social cognitive theory (Bandura, 1994), which together provided a complementary framework for addressing the psychosocial and behavioral challenges experienced by AYLHIV during the transition to adult HIV care.

- Empowerment theory guided the intervention’s focus on strengthening personal agency, participation, and perceived control over health-related decisions, conceptualizing empowerment as a multilevel construct encompassing intrapersonal, interactional, and behavioral domains (Zimmerman, 2000). Within the context of HIV care, this framework supports AYLHIV’s ability to actively engage in health management, navigate healthcare systems, and make informed decisions during a critical developmental period. These principles were operationalized through participatory activities, peer engagement, and structured opportunities for shared reflection and problem-solving.
- Social cognitive theory complemented this approach by informing strategies to promote behavior change through self-efficacy, observational learning, self-regulation, and outcome expectancies (Bandura, 1994). Given the central role of self-efficacy in treatment adherence, health-seeking behaviors, and emotional coping, this framework guided the intervention’s skill-building components. Accordingly, we integrated into the intervention cognitive-behavioral techniques, experiential learning, and peer role modeling to enhance confidence in managing HIV-related challenges and support the adoption and maintenance of adaptive health behaviors.

These theoretical perspectives provided a coherent foundation for EPT, linking individual agency with social learning processes to support sustainable psychosocial and behavioral change within a youth-centered HIV care context.

### Study setting

Participants were recruited from the HIV clinic attached to the Mbarara Regional Referral Hospital (MRRH) in Mbarara city, Uganda. Mbarara city has a population of 264,425 and is the 4th largest city in Uganda by population according to the 2024 census (Uganda Bureau of Statistics, 2024). Most people who access care in the MRRH HIV clinic live in neighboring villages in predominantly rural areas, where livelihoods are based primarily on subsistence agriculture, animal rearing, and small-scale trading challenges of water and food insecurity (Mushavi et al., 2020; Tsai et al., 2011; Tsai et al., 2016). The prevalence of HIV in Mbarara city is estimated at 9.9% more than twice the national prevalence of 5.8% (Ministry of Health, 2021; Uganda AIDS Commission, 2025). The MRRH HIV clinic provides comprehensive HIV care including ART services, viral load monitoring and treatment of opportunistic infections. Although the Ugandan Ministry of Health provides guidelines for AYLHIV transitioning to adult HIV care (Ministry of Health, 2020), little preparation occurs at the study site, due to understaffing. As a result, AYLHIV who are transitioned to adult HIV care often lack the necessary self-management skills to manage their illness and navigate the adult HIV clinic on their own.

## Phase 1: Qualitative Needs assessment

### Study participants

We enrolled three groups of study participants for in-depth interviews: AYLHIV (n=30), caregivers (parents/guardians) of AYLHIV (n=20), and health care providers with experience caring for AYLHIV (n=10). To be eligible for the study, AYLHIV had to be aged 15-24 years, be fully aware of their HIV status, live within 60 km of the clinic, and be able to provide assent or informed consent. Purposive sampling was used to ensure that a diversity of perspectives were represented, including different ages (15–19-year-old adolescents vs. 20–24-year-old young adults, different care statuses (e.g., currently active in care vs. formerly in care), different sexes (boys, girls, men and women), and different stages of transition (in adolescent HIV care vs. adult HIV care). Health care providers were eligible if they were employed at the MRRH HIV clinic (including physicians, counsellors, and/or social workers) and were actively involved in providing health care and related services to ALYHIV. We also included health care providers from the adult HIV care teams to ensure that their views were captured, given their roles in caring for young people with HIV who have transitioned to adult HIV care. We excluded ALYHIV whose HIV status was not fully disclosed to them despite being on antiretroviral therapy (ART); those who were not physically strong enough to remain present for the duration of the interview (e.g., due to physical illness); and those who had difficulty fully understanding the interview questions due to cognitive impairment, after clinical assessment in consultation with a licensed Ugandan psychiatrist. A total of 40 AYLHIV were selected to participate in the intervention.

### Qualitative data collection

Interviews were carried out using a structured guide developed from a review of literature on self-management among AYLHIV, along with input from professionals experienced in HIV care for this group. The guide examined participants’ perspectives on navigating adult HIV care, their self-management skills and needs, the challenges encountered during the transition to adult care, and the role of healthcare providers and informal caregivers in facilitating this process. The guide was originally prepared in English, translated into Runyankore, and subsequently back-translated into English to ensure conceptual equivalence. Data collection was conducted by three graduate-level research assistants in either English or Runyankore, based on participants’ preferences. Each interview lasted approximately one hour, with 15 of the 60 participants opting to be interviewed in Runyankore. All interviews were audio-recorded and then transcribed and translated into English by the research assistants. To ensure accuracy and maintain fidelity to the original content, the transcripts were reviewed by both the research assistants and the first author (SA).

### Qualitative data analysis

Using a thematic analysis approach as outlined by Lochmiller (2021), two members of the research team (SA and PT) independently and iteratively reviewed the first eight transcripts. They exchanged notes, discussed recurring concepts across interviews, and grouped related codes into broader categories to develop a codebook. During this process, emerging themes and sub-themes were identified, with ongoing discussions between the two researchers until consensus was achieved. This iterative approach enabled the researchers to interpret and conceptualize themes related to the self-management needs of AYLHIV as they prepared to transition to adult HIV care. The initial eight transcripts were double-coded to enhance consistency. The remaining transcripts were then divided between the two coders and analyzed independently. Data analysis was supported by MAXQDA software (version 20). Reporting of findings adhered to the Consolidated Criteria for Reporting Qualitative Research (COREQ) guidelines (Booth et al., 2014).

## Phase 2: Intervention Development

### Intervention design process

We followed guidance from the Person-Based Approach and the Medical Research Council (MRC) Framework for developing complex interventions to design an intervention grounded in the needs, perspectives, and context of adolescents and young people living with HIV (AYLHIV) in rural Uganda (Craig et al., 2008; Michie et al., 2011). The Person-Based Approach emphasizes iterative, user-centered development informed by an in-depth understanding of users’ experiences and priorities, which guided our focus on incorporating AYLHIV perspectives throughout the intervention design process (Yardley et al., 2015). The MRC Framework informed the systematic development of the intervention, including identification of key intervention components, engagement of relevant stakeholders, and specification of potential mechanisms of action (Craig et al., 2013). Findings from the qualitative needs assessment were used to identify key psychosocial and behavioral challenges, self-management needs, and transition-related support requirements among AYLHIV. These findings, together with the principles of Empowerment Theory and Social Cognitive Theory, informed the development of an initial intervention outline. The intervention was subsequently refined through stakeholder consultations, expert review, and pretesting to ensure its relevance, acceptability, and feasibility within the local HIV care context.

#### Stakeholder review

To assess the relevance, acceptability, and feasibility of the proposed content, focus group discussions were conducted with key stakeholders, including AYLHIV (n = 10), caregivers of AYLHIV (n = 10), and healthcare providers with experience caring for AYLHIV (n = 10). All participants in these focus group discussions were selected from the 60 individuals who had previously participated in the in-depth interviews. Discussions explored the appropriateness of intervention domains, cultural and social acceptability, session length, delivery strategies, and feasibility within routine HIV care settings. Stakeholders also provided input on the potential of the intervention to motivate, educate, and empower AYLHIV for a successful transition to adult HIV care. Feedback from the focus group discussions was used to refine and strengthen the intervention curriculum prior to expert review.

#### Theory-driven content

To further refine the intervention content, we drew on empowerment theory (Zimmerman, 2000) and social cognitive theory (Bandura, 1994, 2004), which informed the selection of intervention domains and delivery strategies. These frameworks emphasize the importance of personal agency, participation, self-efficacy, and skill development in shaping health behaviors, and guided the inclusion of components aimed at strengthening adolescents’ confidence, knowledge, and capacity to engage in HIV care during the transition to adult services (Bandura, 2004; Benight & Bandura, 2004; Zimmerman, 2000). To ensure transparency in how qualitative findings informed intervention development, we mapped key themes from the in-depth interviews to relevant theoretical constructs and corresponding intervention modules and activities. This mapping illustrates how locally identified needs were integrated with theory to shape the design of the Empowerment and Personal Transformation (EPT) intervention (Supplementary Table 1).

#### Expert review

We facilitated an expert review process to enhance the rigor, clarity, and implementation readiness of the EPT intervention prior to pilot implementation. The review was conducted by three Ugandan clinical psychologists with expertise in adolescent and young adult mental health, psychosocial interventions, and HIV-related psychological care in resource-constrained settings. None of the expert reviewers was a member of the study team or a co-author of this manuscript. The clinical psychologists reviewed the draft intervention manual, module content, and proposed delivery strategies. They focused on assessing the theoretical coherence of the intervention; the appropriateness of content for AYLHIV; and the alignment of activities with principles of empowerment, behavior change, and psychological wellbeing.

Particular attention was given to the emotional and developmental suitability of the modules; sensitivity to HIV-related stigma and disclosure challenges; and the balance between cognitive, emotional, and experiential components. Feedback from the expert reviewers addressed module sequencing, clarity of session objectives, appropriateness of language, and feasibility of delivery by trained facilitators within routine HIV care settings. They also provided guidance on strengthening emotional safety during group sessions, refining experiential and reflective exercises, and ensuring adequate support mechanisms for participants experiencing distress.

All feedback was systematically documented and reviewed by the research team. Revisions were made through an iterative process, resulting in refinements to module structure, simplification of content where necessary, and clearer facilitator guidance. This expert review process contributed to strengthening the content validity, psychological appropriateness, and practical feasibility of the EPT intervention.

### Pretesting

Following expert review and content refinement, the EPT intervention package underwent pretesting to assess its feasibility, clarity, participant engagement, and emotional safety prior to implementation. Pretesting was conducted with 20 AYLHIV, who participated in three group sessions of 10 participants each. These participants met the eligibility criteria for the trial but were excluded from the main trial to avoid contamination. Of the 20 participants involved in pretesting, seven had previously participated in the qualitative interviews used to inform intervention development. The remaining 13 participants were newly recruited from the same HIV clinic. These participants were selected using the same eligibility criteria applied in the main trial and were therefore representative of the target population for whom the intervention was designed. Findings from the qualitative needs assessment, stakeholder consultations, theoretical mapping, expert review, and pretesting informed the final EPT intervention package.

## Phase 3: Pilot Implementation and Feasibility/Acceptability Evaluation

### Pilot participants

A total of 40 AYLHIV were enrolled in the pilot implementation of the EPT intervention. Feasibility and acceptability were evaluated to determine whether the intervention could be delivered as intended and whether it was perceived as relevant, appropriate, and useful for supporting transition to adult HIV care. Participants who completed the intervention were invited to complete the feasibility and acceptability assessment; 37 participants completed these assessments and were included in the analysis. Feasibility assessment focused on the practicality of delivering the intervention within routine HIV care settings, including participant attendance, session completion, session flow, the ability to deliver sessions within the allocated time, and the suitability of the group-based format for AYLHIV. Acceptability assessment focused on participants’ perceptions of the intervention, including the extent to which the intervention was perceived as satisfactory, appealing, and relevant to their transition-related needs. Particular attention was given to participants’ perceptions of the intervention content and delivery approach.

### Study measures

The primary implementation outcomes assessed during pilot implementation of the Empowerment and Personal Transformation (EPT) intervention were acceptability and feasibility. These outcomes were selected based on the implementation outcomes framework developed by Weiner and colleagues (Weiner et al., 2017). Assessments were conducted among AYLHIV enrolled in the pilot implementation of the intervention (n = 40). Of these, 37 participants completed the intervention and the feasibility and acceptability assessment and were included in the analysis. Acceptability was measured using the Acceptability of Intervention Measure (AIM), a four-item scale assessing the extent to which an intervention is perceived as agreeable, satisfactory, and appealing. Items are rated on a five-point Likert scale ranging from strongly disagree (1) to strongly agree (5). Item scores were summed to generate a total score ranging from 4 to 20, with higher scores indicating greater perceived acceptability of the intervention. Feasibility was measured using the Feasibility of Intervention Measure (FIM), a four-item scale assessing the perceived practicality and ease of implementing the intervention as delivered. Items are rated on the same five-point Likert scale. Item scores were summed to produce a total score ranging from 4 to 20, with higher scores indicating greater perceived feasibility of the intervention.

### Data analysis

Quantitative data were analyzed using Stata version 17 (StataCorp, College Station, TX, USA). Analyses were restricted to participants who completed the feasibility and acceptability assessment following participation in the EPT intervention (n = 37). Descriptive statistics were used to summarize participant characteristics and implementation outcomes. Continuous variables were summarized using means and standard deviations, while categorical variables were summarized using frequencies and percentages. Feasibility and acceptability of the Empowerment and Personal Transformation (EPT) intervention were assessed using the Feasibility of Intervention Measure (FIM) and the Acceptability of Intervention Measure (AIM), respectively. Item-level response distributions and total scale scores were examined to assess participants’ perceptions of the intervention. Mean scores and standard deviations were calculated for each measure, and the distribution of responses across the five-point Likert scale categories (strongly disagree to strongly agree) was examined to characterize overall patterns of endorsement.

The internal consistency of the AIM and FIM scales was assessed using Cronbach’s alpha coefficients, with values of 0.70 or higher considered indicative of acceptable reliability.

### Ethical considerations

Ethical approval for the study was obtained from the Mbarara University of Science and Technology Research Ethics Committee (MUST-2021-135), and additional clearance was granted by the Uganda National Council for Science and Technology (UNCST; #HS1697ES). Written informed consent was obtained from all participants prior to their inclusion in the study, in line with UNCST guidelines (Uganda National Council for Science and Technology, 2007). Participants were provided with 25,000 Ugandan Shillings (approximately USD 7 at the time of the study) as compensation for transport. Audio recordings were promptly transferred to a password-protected computer accessible only to two members of the research team (SA and PT). To maintain confidentiality, all identifying information was removed from the data prior to analysis.

## RESULTS

A total of 60 individuals participated, including 30 AYLHIV, 20 caregivers, and 10 healthcare providers.

**Table 1:** Characteristics of the participants involved in the qualitative needs assessment (N=60)

| **Variables** | **Mean (SD) or number** | **%** |
| --- | --- | --- |
| **AYLHIV (N=30)** |  |  |
| *Age, years* | 20 (2.2) |  |
| *Age started ART* | 5.7 (4.2) |  |
| *Years on ART* | 15 (3.8) |  |
| *Sex* |  |  |
| Male | 18 | 60 |
| Female | 12 | 40 |
| *Religion* |  |  |
| Christian | 26 | 87 |
| Moslem | 4 | 13 |
| *Marital status* |  |  |
| Single | 29 | 97 |
| Married | 1 | 3 |
| *Education level* |  |  |
| Primary | 2 | 7 |
| Secondary | 15 | 50 |
| Above secondary | 12 | 43 |
| *Transition status* |  |  |
| Not yet transitioned to adult care | 27 | 90 |
| Transitioned to adult care | 3 | 10 |
| *Employment status* |  |  |
| Formal employment | 2 | 6 |
| Informal employment | 8 | 27 |
| Student | 20 | 67 |
| **Caregivers (n=20)** |  |  |
| Age, years | 47 (9.5) |  |
| Number of children | 3.9 (2.1) |  |
| Number of children in HIV care | 1.3 (0.7) |  |
| *Marital status* |  |  |
| Married | 13 | 65 |
| Single/separated/widowed | 7 | 35 |
| *Religion* |  |  |
| Christian | 19 | 95 |
| Moslem | 1 | 5 |
| *Education level* |  |  |
| Primary | 15 | 75 |
| Above primary | 5 | 25 |
| *Employment* |  |  |
| Farmer | 15 | 75 |
| Business | 5 | 25 |
| **Healthcare providers (N=10)** |  |  |
| Age, years | 39 (10) |  |
| Years providing HIV care | 7.2 (4.9) |  |
| *Sex* |  |  |
| Female | 7 | 70 |
| Male | 3 | 30 |
| *Marital status* |  |  |
| Married | 7 | 70 |
| Single | 3 | 30 |
| *Employment designation* |  |  |
| Clinician | 5 | 50 |
| Counselor | 5 | 50 |

### Qualitative findings informing intervention development

Analysis of interviews with AYLHIV, caregivers, and healthcare providers identified six key themes that were considered important for successful transition to adult HIV care. These findings informed the development of the EPT intervention modules.

## Theme 1: Communication, Self-Advocacy, and Disclosure Skills

Participants emphasized that AYLHIV required strong communication skills to successfully navigate adult HIV care. Many described adolescents as needing greater confidence to communicate with healthcare providers, ask questions, seek clarification, disclose concerns, and advocate for themselves. Participants noted that many adolescents had previously relied on caregivers to communicate on their behalf and therefore lacked experience engaging independently with health services.

> “*They have to build skills of how to communicate… they can learn to say ‘No’ or ‘Yes’ in their favor so that they are not harmed.” ––**Healthcare provider***
>
> *“I think the capability they need here is the ability to speak up… in the adult HIV clinic you will not find health care providers persuading you to talk.” –– **Young adult living with HIV***
>
> *“Most of us adolescents at some point we fear; like we fear to talk to the health care providers because we are not used to those doctors for adults, we fear to disclose our health issues and more so discomfort is also there because we are not used to the place**” ––Young adult living with HIV***

Participants also emphasized the importance of HIV status disclosure as a skill that could facilitate support, adherence, and engagement with healthcare services.

> *“They should be confident enough to disclose their health challenges to the health care providers and other trusted people for them to get proper services.”* ––***Young adult living with HIV***

## Theme 2: Confidence, Empowerment, and Personal Agency

Participants highlighted the importance of confidence, self-esteem, independence, and decision-making. Adolescents were described as needing opportunities to express themselves, exercise their rights, and develop the confidence required to navigate adult HIV care independently.

> *“These children should be left alone and exercise their rights… that is how they will gain confidence.” –**Caregiver***
>
> “*Someone who understands that everyone can contract HIV and having it doesn’t mean the end of the world. Someone who understands that when you take your medicine well you can live a happy life, someone who will not judge his/her parents for infecting him/her with HIV.” ––**Young adult living with HIV***

Participants also emphasized empowering adolescents through knowledge and preparation.

> “If you equip these people with enough knowledge, I think that is the best way of empowering them.” ––***Healthcare provider***
>
> *“We need to consider resilience as a life skill and self-esteem mainly because you find the reasons they give not to be sitting in the adult clinic is like these people are judging us… So that scares them but if they are resilient enough to all that stigma they can handle” **––Healthcare provider***

## Theme 3: Stigma, Disclosure Concerns, and Emotional Challenges

Participants described stigma, fear of judgment, discrimination, and disclosure-related anxiety as major challenges for AYLHIV. These experiences were perceived to negatively affect self-esteem, confidence, and willingness to engage in care.

> *“Everybody at home is treating them in that special way which makes them feel bad about themselves.” ––**Caregiver***
>
> *“During my school days, I hid my HIV status fearing my peers’ reactions. Once they knew, I became a target for mockery, taunts, and humiliation. They would point at me saying, ‘Look at her, she is HIV-positive, a walking corpse.’ These cruel words devastated me, affecting my academic performance and self-esteem.” **––Young adult living with HIV***
>
> *Most of us adolescents at some point we fear; like we fear to talk to the health care providers because we are not used to those doctors for adults, we fear to disclose our health issues and more so discomfort is also there because we are not used to the place.” ––Adolescent living with HIV*

Participants stressed the importance of addressing stigma and helping adolescents develop positive self-perceptions and coping strategies.

## Theme 4: Resilience, Coping, and Interpersonal Skills

Participants emphasized resilience, patience, perseverance, emotional regulation, and problem-solving as important life skills for navigating transition-related challenges. They also highlighted the need for interpersonal skills that would enable adolescents to build relationships with providers, peers, and other adults.

> *“Adolescents and the youth themselves should have the determination to change; they should accept the change and be willing to cope with the new environment” ––**Young adult living with HIV***
>
> ***“****We need to consider resilience as a life skill and self-esteem mainly because you find the reasons they give not to be sitting in the adult clinic is like these people are judging us… So that scares them but if they are resilient enough to all that stigma they can handle.”––Health care provider*
>
> *“They need to stay focused; whatever happens as they change whether good or bad, they should embrace it.” ––**Caregiver***

## Theme 5: Self-Management, Self-Care, and Transition Readiness

Participants described successful transition as requiring adolescents to develop self-management skills and take increasing responsibility for their health. These skills included medication adherence, clinic attendance, self-care, nutrition, sexual health, and independent decision-making.

> “*They should know that anytime you get a problem related to your health you should immediately contact health workers; they should not wait for long because the situation may be worse and end up destroying your health… you should take the responsibility of your own health. You should know that it is your life” ––Young adult living with HIV*.
>
> *“Teach them also to be careful with their health… when they want to engage in sex they should use protection.” ––Caregiver*
>
> *“There are those children who still need someone to remind him/her to take the medicine, to be reminded to go to the hospital on the review date surely you cannot transition such a person.” Young adult living with HIV*

## Theme 6: Emotional Distress, Healing, and the Need for Safe Emotional Expression

Participants described carrying substantial emotional burdens related to HIV, including shame, guilt, grief, self-blame, rejection, and unresolved family experiences. Several participants reported suppressing emotions, crying alone, or struggling to process painful experiences associated with stigma, disclosure, family relationships, and living with HIV. Participants also described the benefits of supportive environments where they could openly share experiences, express emotions, and learn from others facing similar challenges.

> *“Of course, those abuses and taunts impact me deeply… when I am alone, I often break down and cry, overwhelmed by shame and guilt about living with HIV.” **–Adolescent living with HIV***
>
> ***“****As adolescents living with HIV, we often have pressing concerns and emotional struggles that we desperately need to share with someone. However, when we try to open up to health care providers, we are often met with rushed interactions. This lack of emotional support and validation can be devastating, leaving us to carry our burdens alone**.” Adolescent living with HIV***

These findings identified key psychosocial and behavioral needs relevant to successful transition to adult HIV care among AYLHIV. To inform intervention development, the themes were mapped to relevant constructs from Empowerment Theory (Zimmerman, 2000) and Social Cognitive Theory (Bandura, 1994, 2004) and translated into corresponding EPT intervention modules and activities. This process ensured that locally identified needs were systematically integrated with theory in the design of the EPT intervention. A detailed mapping of qualitative findings, theoretical constructs, and intervention components is provided in Supplementary Table S1.

### The Final EPT Intervention

The final EPT intervention comprised six sequential modules designed to address the psychosocial and behavioral challenges experienced by (AYLHIV)during the transition to adult HIV care (Table 3). The intervention integrates principles from Empowerment Theory and Social Cognitive Theory to strengthen communication skills, personal agency, emotional wellbeing, resilience, self-management, and transition readiness. The modules are delivered sequentially to facilitate progressive skill development, beginning with communication and empowerment skills and advancing to self-management, resilience, and emotional processing. Module I, Communication for Care Engagement and Relationships, focuses on communication skills, self-advocacy, disclosure, and relationship-building with healthcare providers, caregivers, and peers.

Module II, Empowerment, Mindfulness, and Self-Regulation, promotes self-awareness, emotional regulation, decision-making, and adaptive health behaviors. Module III, Self-Concept, Emotions, and Beliefs, addresses identity, stigma, disclosure, self-esteem, values, and emotional experiences that influence health behaviors. Module IV, Coping with Stress, Trauma, and Building Resilience, focuses on stress management, acceptance, resilience-building, and adaptive coping strategies. Module V, Autonomy, Responsibility, and Self-Management, emphasizes transition readiness, medication adherence, independent decision-making, sexual and reproductive health, financial literacy, and personal responsibility for health. Module VI, Experiential and Expressive Approaches to Emotional Processing, uses guided reflection, dialogue, and creative expression to facilitate emotional processing, acceptance, healing, and perspective change. Together, the six modules provide a comprehensive, youth-centered, and culturally responsive intervention that combines psychoeducation, skills development, reflection, peer learning, and experiential activities to support successful transition and sustained engagement in adult HIV care.

**Table 2:**
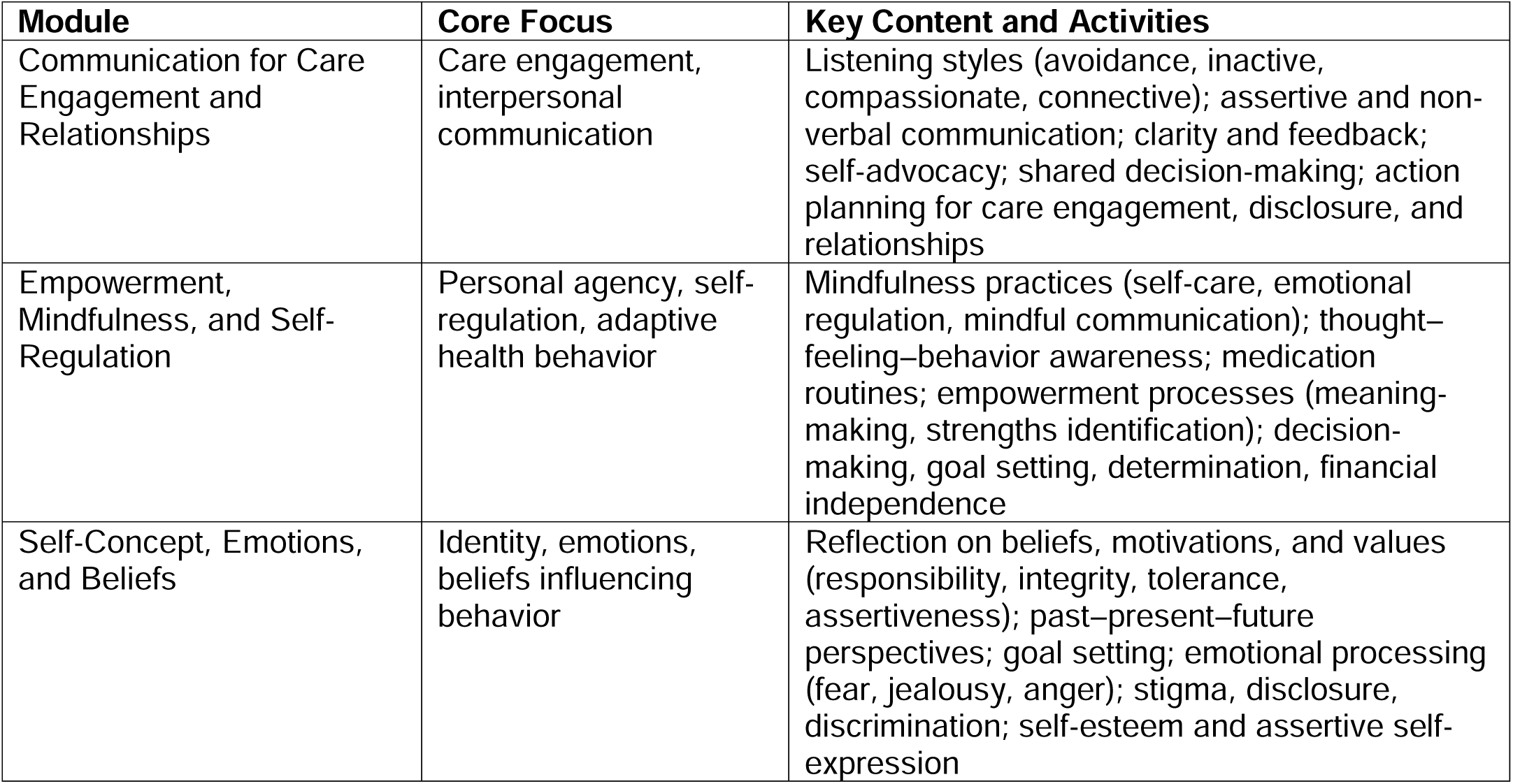
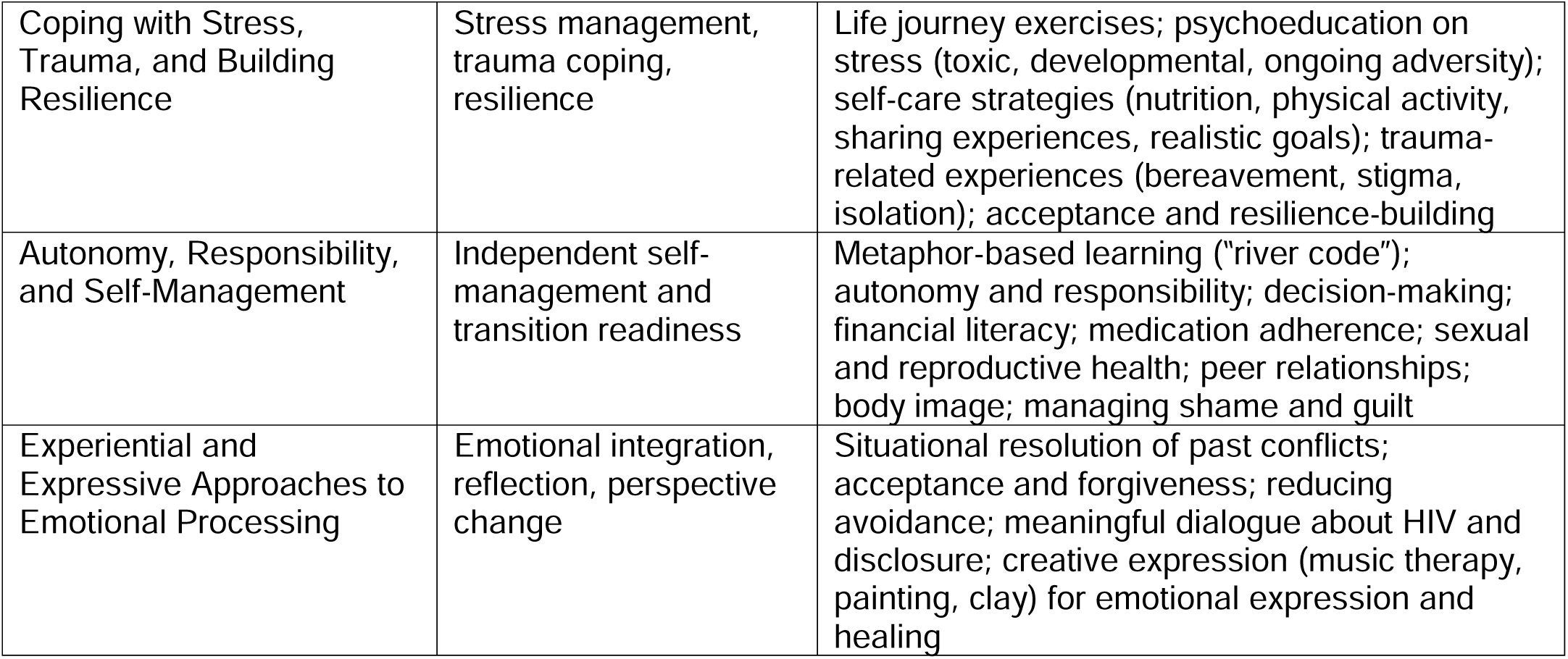
Empowerment and Personal Transformation intervention modules.

## Pilot Implementation and Evaluation

### Participant Characteristics

Characteristics of the 40 AYLHIV enrolled in the pilot implementation of the EPT) intervention are presented in Table 3. Participants had initiated a ART at a mean age of 4.7 years (SD = 5.1) and had been on ART for an average of 14.8 years (SD = 4.7). Slightly more than half of the participants were female (55.0%), and most were enrolled in secondary (55.0%) or tertiary education (27.5%). The intervention was delivered in a group-based format using the six-module EPT curriculum. Of the 40 participants enrolled in the intervention, 37 (92.5%) completed all intervention sessions and contributed to the feasibility and acceptability assessment.

**Table 3.** Characteristics of AYLHIV enrolled in the pilot implementation of the EPT intervention (N = 40)

| Variable | N or Mean (SD) | Percent (%) |
| --- | --- | --- |
| Age at ART initiation (years) | 4.7 ± 5.1 |  |
| Years on ART | 14.8 ± 4.7 |  |
| Sex |  |  |
| Female | 22 | 55 |
| Male | 18 | 45 |
| Education Level |  | 55. |
| Secondary | 22 | 55.0 |
| Tertiary | 11 | 27.5 |
| Primary | 7 | 17.5 |
| Marital Status |  |  |
| Single | 40 | 100 |
| Employment Status |  |  |
| Student | 27 | 67.5 |
| Casual work | 12 | 30.0 |
| Self-employed | 1 | 2.5 |
| Transition Status |  |  |
| Haven't transitioned | 38 | 95.0 |
| Tried and failed | 2 | 5.0 |
| HIV Care Status |  |  |
| Active in care | 40 | 100 |

### Feasibility and Acceptability of the EPT Intervention

Feasibility and acceptability outcomes for the EPT intervention are presented in Table 4. Among the 37 participants who completed the evaluation, both implementation outcomes were highly rated. The Feasibility of Intervention Measure (FIM) demonstrated good internal consistency (Cronbach’s α = 0.86), while the Acceptability of Intervention Measure (AIM) demonstrated excellent internal consistency (Cronbach’s α = 0.92). The mean feasibility score was 18.37 (SD = 2.10) out of a possible 20, indicating that participants perceived the intervention as practical, implementable, and easy to use. Across individual feasibility items, between 94.6% and 100% of participants agreed or strongly agreed that the intervention was implementable, possible, doable, and easy to use. Thehe mean acceptability score was 18.51 (SD = 2.02) out of a possible 20, suggesting high levels of participant satisfaction with the intervention. More than 94% of participants agreed or strongly agreed that the intervention met their approval, was appealing, and was welcomed.

**Table 4:** Feasibility and acceptability of the EPT Intervention (n = 37)

| Measure | Mean (SD) | Strongly Agree<br>n (%) * | Agree<br>n (%) * |
| --- | --- | --- | --- |
| Feasibility of the intervention (FIM) |  |  |  |
| Total FIM score | 18.37 (2.10) |  |  |
| The intervention is implementable |  | 23 (62.2%) | 14 (37.8%) |
| The intervention seems possible |  | 23 (62.2%) | 12 (32.4%) |
| The intervention seems doable |  | 26 (70.3%) | 9 (24.3%) |
| The intervention seems easy to use |  | 25 (67.6%) | 8 (21.6%) |
| Acceptability of the intervention (AIM) |  |  |  |
| Total AIM score | 18.51 (2.02) |  |  |
| The intervention meets my approval |  | 28 (75.7%) | 9 (24.3%) |
| The intervention is appealing to me |  | 24 (64.9%) | 11 (29.7%) |
| I like the intervention |  | 22 (59.5%) | 13 (35.1%) |
| I welcome the intervention |  | 25 (67.6%) | 10 (27.0%) |

## Discussion

In this mixed methods study of 60 AYLHIV, caregivers, and health care providers in rural Uganda, we developed an EPT intervention to aid AYLHIV transitioning to adult HIV care. The design process included extensive input from all stakeholders and was refined through pretesting before being deployed in a sample of 40 AYLHIV. We found high levels of feasibility and acceptability, with most participants completing all sessions and either agreeing or strongly agreeing that the intervention was implementable, possible, easy to use, and appealing.

High feasibility scores indicate that the intervention was perceived as practical and implementable within the existing HIV care context. This finding is particularly important given that transition interventions often operate within constrained health systems and must be integrated into routine service delivery (Atun et al., 2010). Feasibility is a critical early implementation outcome, as interventions perceived as difficult to deliver or incompatible with existing systems are unlikely to be adopted or sustained (Powell et al., 2019; Proctor et al., 2023). This consideration is particularly relevant in the context of transition from adolescent to adult HIV care, which is a complex process extending beyond clinical transfer to encompass developmental, psychosocial, and structural changes (Ashaba et al., 2023; Chew et al., 2025). During the transition period, AYLHIV are expected to assume increasing responsibility for their health while simultaneously navigating identity development, social relationships, and growing independence (Hussen et al., 2017; Tanner et al., 2016). Within this context, the high feasibility observed in this study suggests that empowerment-focused approaches can be delivered in a manner that is responsive to these intersecting demands, without imposing excessive burden on the AYLHIV or health care providers (Stepanian et al., 2023; Tanner et al., 2024).

Acceptability of the intervention was also high, indicating strong participant approval and perceived relevance. Acceptability is particularly salient for empowerment and personal transformation interventions, which depend on participant engagement, openness, and sustained involvement (Sekhon et al., 2017). This is especially important for AYLHIV, who often experience stigma, uncertainty about adulthood, and anxiety related to disclosure and long-term health, factors that can shape how they engage with transition-related interventions (Ashaba et al., 2023; Dahourou et al., 2017). As such, the positive reception of the intervention suggests that it aligned closely with the lived experiences and psychosocial needs of the participants.

Consistent with previous research, youth-centered, strengths-based, and participatory approaches tend to be more acceptable to AYLHIV than interventions that focus narrowly on adherence to medicines or clinical compliance (Chew et al., 2024; Embleton et al., 2024). By emphasizing empowerment and personal transformation, the intervention may have fostered a sense of ownership, relevance, and personal meaning among participants (Mouchrek & Benson, 2023).

Successful transition to adult HIV care requires more than continuity of clinical services and requires readiness to manage one’s health independently (Mbalinda et al., 2021; Shimbre et al., 2024; Zanoni et al., 2024). Empowerment-based interventions are well positioned to support this process by strengthening self-efficacy, confidence, and decision-making capacity (Mutumba et al., 2019). The high acceptability observed in this study suggests that participants perceived the intervention as supportive of their growing autonomy and preparedness for adult care.

Developmental theory underscores late adolescence and young adulthood as critical periods for identity formation and consolidation of self-management skills (Lanyon et al., 2020; Machado et al., 2016). In the context of HIV, empowerment has been associated with improved engagement in care, better self-management, and enhanced psychosocial well-being (Ashaba et al., 2024; Enane et al., 2018).The positive reception of the intervention in this study suggests that it may enhance transition readiness by addressing these underlying psychosocial dimensions alongside clinical care (Shimbre et al., 2024). The minimal neutral responses and near absence of disagreement across feasibility and acceptability items further strengthen the interpretation of these findings. Neutral responses may indicate ambivalence, lack of clarity, or limited perceived relevance, all of which can undermine intervention uptake (Rosler et al., 2024). The strong endorsement across items suggests a high degree of perceived relevance and clarity among participants, an encouraging indicator given the heterogeneity of AYLHIV with respect to age, social context, and duration of HIV diagnosis.

However, these findings should be interpreted with caution due to the potential for social desirability bias. Participants may have felt inclined to provide favorable responses to an intervention delivered within a clinical setting by individuals thought to be associated with their HIV care, a phenomenon well documented in health research (Grimm, 2010; Krumpal, 2013).

Although feasibility and acceptability assessments were conducted in the absence of facilitators by research assistants who were not involved in delivering the intervention, responses may still have been influenced by perceived expectations or a desire to please healthcare providers, potentially leading us to overestimate the intervention’s feasibility and acceptability. In addition, participants were recruited from a single regional referral hospital clinic and were largely engaged in HIV care and awaiting transition to adult services, which may limit the generalizability of the findings to other populations of AYLHIV and healthcare settings The EPT intervention may be less feasible or acceptable among AYLHIV who are intermittently or poorly engaged in care. As such, the intervention is likely to be most applicable to AYLHIV who are already linked to care and preparing for transition to adult services. Future research should assess the adaptability and effectiveness of the intervention across more diverse populations and settings to enhance its generalizability. Despite these limitations, the high levels of feasibility and acceptability observed provide important preliminary evidence supporting the potential of the EPT intervention as a promising, contextually relevant approach to improving transition readiness among AYLHIV in routine HIV care settings.

## Conclusions

Our findings contribute to the growing evidence base supporting the integration of empowerment-focused approaches into HIV transition programming (Enane et al., 2020). While many transition models emphasize structural coordination between pediatric and adult services, fewer explicitly address the personal and psychosocial transformation required of young people during this period (Lanyon et al., 2020; Machado et al., 2016). Interventions that are both feasible and acceptable from the perspective of AYLHIV may enhance existing transition frameworks by strengthening internal capacities such as self-confidence, self-advocacy, and future orientation (Ahmed et al., 2023; Ferry et al., 2022). Given persistent gaps in retention and viral suppression among young people following transition to adult HIV care, particularly in resource-limited settings, scalable and youth-acceptable empowerment interventions may play a critical complementary role in improving long-term outcomes.

## Data Availability

All data produced in the present work are contained in the manuscript

## Acknowledgements

We would like to express our gratitude to the study participants who volunteered their time to participate and without whom this study would not have been feasible. This research was funded by the Fogarty International Center of the U.S. National Institutes of Health (NIH) under award number K43TW011929. ACT acknowledges salary support from NIH K24DA061696. The content is solely the responsibility of the authors and does not necessarily represent the official views of the NIH.

## Ethics approval

The study was approved by the Research Ethics Committee of the Mbarara University of Science and Technology (#20/08-19) and the Partners Human Research Committee (#2019P003451). The study also received clearance from the Uganda National Council for Science and Technology (#HS512ES)

## Consent to participate

All Participants provided written informed assent and /or consent to participate in the study.

## Consent for publication

Participants consented to having their data published.

## Availability of data and material

Not applicable

## Code availability

Not applicable

## Author contributions

SA was responsible for conceptualizing the study including development of the methodology, secured funding, provided leadership, and wrote the original draft and oversaw revisions of the manuscript. PT was responsible for data collection and revised the final manuscript. CB was responsible for daily project administration and supervision of research and revised the final version of the manuscript. DN, AC, and JMK reviewed and edited the final version of the manuscript. SM supervised and guided the data collection procedures, and reviewed the final version of the manuscript. BCZ guided the data analysis process, and revised the final version of the manuscript. ACT provided leadership and guidance during conceptualization of the study, data analysis, and contributed to the original draft and revised the final version of the manuscript.

## References

1. Aantjes, C. J., Ramerman, L., & Bunders, J. F. (2014). A systematic review of the literature on self-management interventions and discussion of their potential relevance for people living with HIV in sub-Saharan Africa. Patient education and counseling, 95(2), 185–200.

2. Acuña Mora, M., Sparud-Lundin, C., Bratt, E.-L., & Moons, P. (2019). Empowering young persons during the transition to adulthood. In Transition from pediatric to adult healthcare services for adolescents and young adults with long-term conditions: An international perspective on nurses’ roles and interventions (pp. 19–46). Springer.

3. Ahmed, C. V., Doyle, R., Gallagher, D., Imoohi, O., Ofoegbu, U., Wright, R., Yore, M. A., Brooks, M. J., Flores, D. D., & Lowenthal, E. D. (2023). A systematic review of peer support interventions for adolescents living with HIV in sub-Saharan Africa. AIDS patient care and STDs, 37(11), 535–559.

4. Akama, E. O., Beres, L. K., Kulzer, J. L., Ontuga, G., Adhiambo, H., Bushuru, S., Nyagesoa, E., Osoro, J., Opondo, I., & Sang, N. (2023). A youth-centred approach to improving engagement in HIV services: human-centred design methods and outcomes in a research trial in Kisumu County, Kenya. BMJ Global Health, 8(11).

5. Areri, H. A., Marshall, A., & Harvey, G. (2022). Self-efficacy for self-management and its influencing factors among adults living with HIV on antiretroviral therapy in northwest Ethiopia. AIDS care, 34(12), 1595–1601.

6. Ashaba, S., Baguma, C., Tushemereirwe, P., Nansera, D., Maling, S., Tsai, A. C., & Zanoni, B. C. (2024). A qualitative analysis of self-management needs of adolescents and young adults living with perinatally acquired HIV in rural, southwestern Uganda. PLOS Global Public Health, 4(3), e0003037.

7. Ashaba, S., Zanoni, B. C., Baguma, C., Tushemereirwe, P., Nuwagaba, G., Kirabira, J., Nansera, D., Maling, S., & Tsai, A. C. (2023). Challenges and fears of adolescents and young adults living with HIV facing transition to adult HIV care. AIDS and Behavior, 27(4), 1189–1198.

8. Ashaba, S., Zanoni, B. C., Baguma, C., Tushemereirwe, P., Nuwagaba, G., Nansera, D., Maling, S., & Tsai, A. C. (2022). Perspectives About Transition Readiness Among Adolescents and Young People Living With Perinatally Acquired HIV in Rural, Southwestern Uganda: A Qualitative Study. Journal of the Association of Nurses in AIDS Care, 33(6), 613–623.

9. Atun, R., de Jongh, T., Secci, F., Ohiri, K., & Adeyi, O. (2010). A systematic review of the evidence on integration of targeted health interventions into health systems. Health policy and planning, 25(1), 1–14.

10. Auld, A. F., Agolory, S. G., Shiraishi, R. W., Wabwire-Mangen, F., Kwesigabo, G., Mulenga, M., Hachizovu, S., Asadu, E., Tuho, M. Z., & Ettiegne-Traore, V. (2014). Antiretroviral therapy enrollment characteristics and outcomes among HIV-infected adolescents and young adults compared with older adults––seven African countries, 2004-2013. MMWR. Morbidity and mortality weekly report, 63(47), 1097–1103.

11. Bandura, A. (1994). Social cognitive theory and exercise of control over HIV infection. In Preventing AIDS: Theories and methods of behavioral interventions (pp. 25-59). Springer.

12. Bandura, A. (2004). Health promotion by social cognitive means. Health education & behavior, 31(2), 143–164.

13. Benight, C. C., & Bandura, A. (2004). Social cognitive theory of posttraumatic recovery: The role of perceived self-efficacy. Behaviour research and therapy, 42(10), 1129–1148.

14. Booth, A., Hannes, K., Harden, A., Noyes, J., Harris, J., & Tong, A. (2014). COREQ (consolidated criteria for reporting qualitative studies). Guidelines for reporting health research: a user’s manual, 214–226.

15. Cervia, J. S. (2013). Easing the transition of HIV-infected adolescents to adult care. AIDS Patient Care and STDs, 27(12), 692–696.

16. Chew, H., Ahonkhai, A., Ivey, C., Desai, N., & Zanoni, B. (2025). Transition from adolescent to adult care for young people living with HIV: a systematic review of needs, barriers, and interventions. Journal of Adolescent Health.

17. Chew, H., Bonnet, K., Schlundt, D., Hill, N., Pierce, L., Ahonkhai, A., & Desai, N. (2024). Mixed methods evaluation of a youth-friendly clinic for young people living with HIV transitioning from pediatric care. Tropical Medicine and Infectious Disease, 9(9), 198.

18. Craig, P., Dieppe, P., Macintyre, S., Michie, S., Nazareth, I., & Petticrew, M. (2008). Developing and evaluating complex interventions: the new Medical Research Council guidance. BMJ, 337, a1655.

19. Craig, P., Dieppe, P., Macintyre, S., Michie, S., Nazareth, I., & Petticrew, M. (2013). Developing and evaluating complex interventions: the new Medical Research Council guidance. International journal of nursing studies, 50(5), 587–592.

20. Crowley, T., van der Merwe, A., & Skinner, D. (2019, Jul-Aug). Adolescent HIV Self-management: Lived Experiences of Adolescents, Caregivers, and Health Care Workers in a South African Context. J Assoc Nurses AIDS Care, 30(4), e7–e19. 10.1097/jnc.0000000000000098

21. Dahourou, D. L., Gautier-Lafaye, C., Teasdale, C. A., Renner, L., Yotebieng, M., Desmonde, S., Ayaya, S., Davies, M. A., & Leroy, V. (2017). Transition from paediatric to adult care of adolescents living with HIV in sub-Saharan Africa: challenges, youth-friendly models, and outcomes. Journal of the International AIDS Society, 20, 21528.

22. Davies, M. A., Tsondai, P., Tiffin, N., Eley, B., Rabie, H., Euvrard, J., Orrell, C., Prozesky, H., Wood, R., & Cogill, D. (2017). Where do HIV-infected adolescents go after transfer?–Tracking transition/transfer of HIV-infected adolescents using linkage of cohort data to a health information system platform. Journal of the International AIDS Society, 20, 21668.

23. Embleton, L., Boal, A., Sawarkar, S., Chory, A., Bandanapudi, R. M., Patel, T., Levinson, C., Vreeman, R., Wu, W.-J., & Diaz, A. (2024). Characterizing models of adolescent and youth-friendly health services in sub-Saharan Africa: a scoping review. International journal of adolescent medicine and health, 36(3), 203–236.

24. Enane, L. A., Apondi, E., Toromo, J., Bosma, C., Ngeresa, A., Nyandiko, W., & Vreeman, R. C. (2020). “A problem shared is half solved”–a qualitative assessment of barriers and facilitators to adolescent retention in HIV care in western Kenya. AIDS care, 32(1), 104–112.

25. Enane, L. A., Vreeman, R. C., & Foster, C. (2018). Retention and adherence: global challenges for the long-term care of adolescents and young adults living with HIV. Current Opinion in HIV and AIDS, 13(3), 212–219.

26. Fair, C. D., Sullivan, K., & Gatto, A. (2010). Best practices in transitioning youth with HIV: perspectives of pediatric and adult infectious disease care providers. *Psychology*, Health & Medicine, 15(5), 515–527.

27. Ferry, A., Seeley, J., Weiss, H. A., & Simms, V. (2022). Health-related needs reported by adolescents living with HIV and receiving antiretroviral therapy in sub-Saharan Africa: a systematic literature review. Journal of the International AIDS Society, 25(8), e25921.

28. Garofalo, R., Kuhns, L. M., Reisner, S. L., Biello, K., & Mimiaga, M. J. (2018). Efficacy of an empowerment-based, group-delivered HIV prevention intervention for young transgender women: the project LifeSkills randomized clinical trial. JAMA pediatrics, 172(10), 916–923.

29. Grimm, P. (2010). Social desirability bias. Wiley international encyclopedia of marketing.

30. Hussen, S. A., Chakraborty, R., Knezevic, A., Camacho-Gonzalez, A., Huang, E., Stephenson, R., & Del Rio, C. (2017). Transitioning young adults from paediatric to adult care and the HIV care continuum in Atlanta, Georgia, USA: a retrospective cohort study. Journal of the International AIDS Society, 20(1), 21848.

31. Jao, J., Fairlie, L., Griffith, D., & Agwu, A. L. (2016). The Challenge of and Opportunities for Transitioning and Maintaining a Continuum of Care Among Adolescents and Young Adults Living with HIV in Resource Limited Settings. Current tropical medicine reports, 3(4), 149.

32. Krumpal, I. (2013). Determinants of social desirability bias in sensitive surveys: a literature review. Quality & quantity, 47(4), 2025–2047.

33. Kuhns, L. M., Mimiaga, M. J., Reisner, S. L., Biello, K., & Garofalo, R. (2017). Project LifeSkills-a randomized controlled efficacy trial of a culturally tailored, empowerment-based, and group-delivered HIV prevention intervention for young transgender women: study protocol. BMC public health, 17, 713.

34. Lanyon, C., Seeley, J., Namukwaya, S., Musiime, V., Paparini, S., Nakyambadde, H., Matama, C., Turkova, A., & Bernays, S. (2020). “Because we all have to grow up”: supporting adolescents in Uganda to develop core competencies to transition towards managing their HIV more independently. Journal of the International AIDS Society, 23, e25552.

35. Lochmiller, C. R. (2021). Conducting thematic analysis with qualitative data. The Qualitative Report, 26(6), 2029–2044.

36. Machado, D. M., Galano, E., de Menezes Succi, R. C., Vieira, C. M., & Turato, E. R. (2016). Adolescents growing with HIV/AIDS: experiences of the transition from pediatrics to adult care. The Brazilian Journal of Infectious Diseases, 20(3), 229–234.

37. MacPherson, P., Munthali, C., Ferguson, J., Armstrong, A., Kranzer, K., Ferrand, R. A., & Ross, D. A. (2015). Service delivery interventions to improve adolescents’ linkage, retention and adherence to antiretroviral therapy and HIV care. Tropical medicine & international health, 20(8), 1015–1032.

38. Mbalinda, S. N., Bakeera-Kitaka, S., Lusota, D. A., Musoke, P., Nyashanu, M., & Kaye, D. K. (2021). Transition to adult care: Exploring factors associated with transition readiness among adolescents and young people in adolescent ART clinics in Uganda. PLoS One, 16(4), e0249971.

39. Mburu, G., Ram, M., Oxenham, D., Haamujompa, C., Iorpenda, K., & Ferguson, L. (2014). Responding to adolescents living with HIV in Zambia: a social–ecological approach. Children and Youth Services Review, 45, 9–17.

40. McDonagh, J., & Kelly, D. (2010). The challenges and opportunities for transitional care research. Pediatric transplantation, 14(6), 688–700.

41. Michie, S., Van Stralen, M. M., & West, R. (2011). The behaviour change wheel: a new method for characterising and designing behaviour change interventions. Implementation science, 6(1), 42.

42. Ministry of Health. (2020). Consolidated guidelines for the prevention and treatment of HIV and AIDS in Uganda. https://differentiatedservicedelivery.org

43. Ministry of Health. (2021). Facts on HIV and AIDS in Uganda based on data ending December 2020. [Record #154 is using a reference type undefined in this output style.]

44. Mouchrek, N., & Benson, M. (2023). The theory of integrated empowerment in the transition to adulthood: concepts and measures. Frontiers in sociology, 8, 893898.

45. Mulawa, M. I., Knippler, E. T., Al-Mujtaba, M., Wilkinson, T. H., Ravi, V. K., & Ledbetter, L. S. (2023). Interventions to improve adolescent HIV care outcomes. Current HIV/AIDS Reports, 20(4), 218–230.

46. Mushavi, R. C., Burns, B. F., Kakuhikire, B., Owembabazi, M., Vořechovská, D., McDonough, A. Q., Cooper-Vince, C. E., Baguma, C., Rasmussen, J. D., & Bangsberg, D. R. (2020). “When you have no water, it means you have no peace”: a mixed-methods, whole-population study of water insecurity and depression in rural Uganda. Social Science & Medicine, 245, 112561.

47. Mutumba, M., Mugerwa, H., Musiime, V., Gautam, A., Nakyambadde, H., Matama, C., & Stephenson, R. (2019). Perceptions of strategies and intervention approaches for HIV self-management among Ugandan adolescents: A qualitative study. Journal of the International Association of Providers of AIDS Care (JIAPAC), 18, 2325958218823246.

48. Ness, T. E., Agrawal, V., Guffey, D., Small, A., Simelane, T., Dlamini, S., Petrus, J., & Lukhele, B. (2021). Impact of using creative arts programming to support HIV treatment in adolescents and young adults in Eswatini. AIDS Research and Therapy, 18(1), 100.

49. Powell, B. J., Fernandez, M. E., Williams, N. J., Aarons, G. A., Beidas, R. S., Lewis, C. C., McHugh, S. M., & Weiner, B. J. (2019). Enhancing the impact of implementation strategies in healthcare: a research agenda. Frontiers in public health, 7, 3.

50. Proctor, E. K., Bunger, A. C., Lengnick-Hall, R., Gerke, D. R., Martin, J. K., Phillips, R. J., & Swanson, J. C. (2023). Ten years of implementation outcomes research: a scoping review. Implementation Science, 18(1), 31.

51. Rosler, N., Wiener-Blotner, O., Micheles, O. H., & Sharvit, K. (2024). Understanding Reactions to Informative Process Model Interventions: Ambivalence as a Mechanism of Change. *Behavioral sciences (Basel*, Switzerland), 14(12), 1152.

52. Schwartz, L., Tuchman, L., Hobbie, W., & Ginsberg, J. (2011). A social-ecological model of readiness for transition to adult-oriented care for adolescents and young adults with chronic health conditions. Child: care, health and development, 37(6), 883–895.

53. Sekhon, M., Cartwright, M., & Francis, J. J. (2017). Acceptability of healthcare interventions: an overview of reviews and development of a theoretical framework. BMC Health Services Research, 17(1), 88.

54. Sell, K., Amella, E., Mueller, M., Andrews, J., & Wachs, J. (2016). Use of social cognitive theory to assess salient clinical research in chronic disease self-management for older adults: An integrative review. Open Journal of Nursing, 6(3), 213–228.

55. Shimbre, M. S., Abay, G., Belete, A. G., Mengesha, M. M., & Ma, W. (2024). Predictors of successful transition of adolescents and young adults living with HIV from pediatric to adult-oriented care in southern Ethiopia: a retrospective cohort study. BMC Health Services Research, 24(1), 836.

56. Slogrove, A. L., Mahy, M., Armstrong, A., & Davies, M. A. (2017). Living and dying to be counted: What we know about the epidemiology of the global adolescent HIV epidemic. Journal of the International AIDS Society, 20, 21520.

57. Stepanian, N., Larsen, M. H., Mendelsohn, J. B., Mariussen, K. L., & Heggdal, K. (2023). Empowerment interventions designed for persons living with chronic disease-a systematic review and meta-analysis of the components and efficacy of format on patient-reported outcomes. BMC Health Services Research, 23(1), 911.

58. Tanner, A. E., Mertus, S., Jibriel, M. S. E., Urquhart, R., Phillips, K., Dowshen, N., Dutta, S., Goldstein, M. H., Lee, S., & Knowles, K. (2024). Transitioning adolescents to adult HIV care in the United States: implementation lessons from the iTransition Intervention Pilot Trial. Tropical Medicine and Infectious Disease, 9(12), 297.

59. Tanner, A. E., Philbin, M. M., DuVal, A., Ellen, J., Kapogiannis, B., Fortenberry, J. D., & Interventions, A. T. N. f. H. A. (2016). Transitioning HIV-positive adolescents to adult care: lessons learned from twelve adolescent medicine clinics. Journal of pediatric nursing, 31(5), 537–543.

60. Tsai, A. C., Bangsberg, D. R., Emenyonu, N., Senkungu, J. K., Martin, J. N., & Weiser, S. D. (2011). The social context of food insecurity among persons living with HIV/AIDS in rural Uganda. Social science & medicine, 73(12), 1717–1724.

61. Tsai, A. C., Kakuhikire, B., Mushavi, R., Vořechovská, D., Perkins, J. M., McDonough, A. Q., & Bangsberg, D. R. (2016). Population-based study of intra-household gender differences in water insecurity: reliability and validity of a survey instrument for use in rural Uganda. Journal of Water and Health, 14(2), 280–292.

62. Uganda AIDS Commission. (2025). Uganda HIV and AIDS factsheet based on data ending 31st December 2024.

63. Uganda Bureau of Statistics. (2024). Main Cities by population in Uganda.

64. Weiner, B. J., Lewis, C. C., Stanick, C., Powell, B. J., Dorsey, C. N., Clary, A. S., Boynton, M. H., & Halko, H. (2017). Psychometric assessment of three newly developed implementation outcome measures. Implementation science, 12(1), 108.

65. Yardley, L., Morrison, L., Bradbury, K., & Muller, I. (2015). The person-based approach to intervention development: application to digital health-related behavior change interventions. Journal of medical Internet research, 17(1), e4055.

66. Zanoni, B. C., Archary, M., Sibaya, T., Musinguzi, N., Gethers, C. T., Goldstein, M., Bergam, S., Psaros, C., Marconi, V. C., & Haberer, J. E. (2024). Acceptability, feasibility and preliminary effectiveness of the mHealth intervention, InTSHA, on retention in care and viral suppression among adolescents with HIV in South Africa: A pilot randomized clinical trial. AIDS care, 36(7), 983–992.

67. Zimmerman, M. A. (2000). Empowerment theory: Psychological, organizational and community levels of analysis. In Handbook of community psychology (pp. 43–63). Springer.

