## supplementary Table 1 for "Development and pilot testing of an empowerment-based intervention for adolescents and young adults living with HIV transitioning to adult HIV care in Uganda"

**Supplementary Table: Mapping qualitative findings and theoretical constructs to EPT modules and activities**

| **Qualitative theme / finding** | Need identified from qualitative data | Relevant theoretical construct(s) | Corresponding EPT module | How this informed intervention content / activities |
| --- | --- | --- | --- | --- |
| Communication difficulties with providers, caregivers, and others | Adolescents were described as shy, fearful, and often unable to ask questions, express concerns, or communicate effectively in care and social settings. | Self-efficacy; behavioral capability; self-advocacy; participation | Communication for Care Engagement and Relationships | Informed content on assertive communication, self-advocacy, non-verbal communication, feedback, shared decision-making, and action planning for care engagement, disclosure, and relationships. |
| Need for confidence, personal agency, and self-regulation | Participants emphasized the importance of confidence, self-belief, decision-making, and emotional regulation in helping adolescents manage transition and care. | Empowerment; self-efficacy; agency; self-regulation | Empowerment, Mindfulness, and Self-Regulation | Guided inclusion of mindfulness practices, awareness of thoughts–feelings–behaviors, empowerment exercises, strengths identification, goal setting, decision-making, and routines for medication-taking and self-care. |
| Stigma, disclosure concerns, low self-esteem, and emotional struggles | Adolescents described stigma, fear of disclosure, shame, judgment, and internal struggles related to identity and HIV status. | Empowerment; self-efficacy; identity; cognitive appraisal | Self-Concept, Emotions, and Beliefs | Informed activities addressing stigma, disclosure, self-esteem, assertive self-expression, emotional awareness, beliefs and values, and reflection on identity, goals, and future orientation. |
| Stress, trauma, adversity, and need for resilience | Participants described the emotional burden of HIV, stigma, isolation, bereavement, and difficult life experiences, and emphasized the need for resilience and acceptance. | Resilience; coping self-efficacy; adaptive coping | Coping with Stress, Trauma, and Building Resilience | Informed psychoeducation on stress and adversity, life journey reflection, acceptance, resilience-building, and self-care strategies such as sharing experiences, realistic goal setting, and emotional coping. |
| Limited independence and self-management for adult HIV care | Adolescents were seen as needing stronger skills in medication adherence, clinic attendance, independent decision-making, and responsibility for their care. | Behavioral capability; self-management; autonomy; personal agency | Autonomy, Responsibility, and Self-Management | Informed content focused on transition readiness, autonomy, decision-making, medication adherence, responsibility, financial literacy, sexual and reproductive health, peer relationships, and self-management in daily life. |
| Need for emotional expression, healing, and processing difficult experiences | Some findings pointed to unresolved emotional pain, fear, silence, and avoidance, suggesting the need for safe spaces to process emotions and lived experiences. | Emotional processing; self-reflection; empowerment; coping | Experiential and Expressive Approaches to Emotional Processing | Informed use of expressive and reflective activities such as meaningful dialogue, conflict resolution, acceptance, forgiveness, and creative approaches (e.g., music, painting, clay) to support emotional expression and healing. |
| Fear and uncertainty about transition to adult HIV care | Adolescents expressed anxiety about moving into adult HIV care and wanted orientation, preparation, and support to understand what to expect. | Self-efficacy; readiness; empowerment | Autonomy, Responsibility, and Self-Management; Communication for Care Engagement and Relationships | Supported inclusion of transition readiness activities, practical preparation for adult care, and communication strategies to help adolescents ask questions, seek clarification, and navigate new care systems. |
| Need for peer support and shared experience | Adolescents valued support from peers and preferred transitioning with others who had similar experiences. | Observational learning; social modeling; collective empowerment | Communication for Care Engagement and Relationships; Coping with Stress, Trauma, and Building Resilience | Supported inclusion of peer-based interaction, shared reflection, and activities that allow adolescents to learn from others’ experiences and reduce isolation during transition. |
| Need for caregiver and provider support during transition | Participants emphasized that transition required continued involvement of caregivers and supportive health care providers. | Social support; environmental influences; participation | Cross-cutting across all modules | Supported the design of intervention content that recognizes transition as a relational process and encourages adolescents to identify, communicate with, and draw support from caregivers, providers, and other trusted adults. |
| Need for HIV and transition-related information | Participants wanted information about ART, adult clinic procedures, treatment expectations, retention in care, and benefits of adult care. | Knowledge; behavioral capability | Cross-cutting, especially Autonomy, Responsibility, and Self-Management and Empowerment, Mindfulness, and Self-Regulation | Informed psychoeducational content embedded across modules to improve knowledge, readiness, and confidence in managing HIV care during transition. |

**EPT = Empowerment and Personal Transformation intervention

This table maps key qualitative themes to theoretical constructs and corresponding EPT modules and activities.
